# Mesoamerican Nephropathy in Central Panama

**DOI:** 10.1101/2022.02.19.22271236

**Authors:** Karen Courville, Norman Bustamante, Bárbara Hurtado, Maydelin Pecchio, Clarissa Rodríguez, Virginia Núñez-Samudio, Iván Landires

## Abstract

**Background:** In the last three decades, there has been an increase in the frequency of patients diagnosed with chronic kidney disease of nontraditional causes (CKDnt) in the Mesoamerican region. A region with an increased frequency of patients with chronic kidney disease (CKD) has been identified in central Panama. The present study aims to characterize the clinical presentation of patients with CKDnt in an understudied population of the central region of Panama and compare them with patients with traditional CKD (CKDt).

**Methods:** A retrospective descriptive study was conducted in a nephrology reference hospital in the central provinces of Herrera and Los Santos, comparing a group of 15 patients with CKDnt to 91 patients with CKDt. Sociodemographic variables, personal history, laboratory parameters, and of renal ultrasound were compared.

**Results:** CKDnt is more common among relatively younger male patients who engage in strenuous work activities at high temperatures. CKDnt is not associated with type 2 diabetes mellitus or chronic hypertension, as it is the case in patients with CKDt. Renal atrophy and hyperuricemia are significant clinical markers of CKDnt.

**Conclusion:** To our knowledge, this is the first study in Panama and one of the few in Central America and the world to address the clinical presentation of patients with CKDnt compared to patients with CKDt. Because CKDnt remains asymptomatic for a long time, early detection is important, and efforts should be directed at halting disease progression at an early stage. Current evidence can also inform policies addressing occupational and environmental risk factors associated with CKDnt.

## Background

Chronic kidney disease (CKD) belongs to the group of noncommunicable diseases and, according to estimates from the Global Burden of Disease group, produced a 19.6% increase in the disability-adjusted life years (DALYs) and has been associated with 4% of deaths worldwide from 2005 to 2015, representing 2.2 million deaths per year [1,2].

In 60% of patients, the cause of CKD is identified. Among the traditional causes of CKD are diabetes mellitus, essential hypertension, and obesity, followed by a minority secondary to immunological diseases, nephrolithiasis, and genetic conditions [3]. Approximately 25% of adult patients with CKD may have a family history of this pathology. In 10% of patients, the cause of CKD cannot be identified, which is classified as unknown or nontraditional cause (CKDnt) [4]. For several years, an increase in the frequency of diagnoses of CKDnt from agricultural areas of Central America (El Salvador, Nicaragua, Guatemala, and Costa Rica) has been identified, which is why it has been denominated Mesoamerican nephropathy. This pathology has been presented mainly in young male patients with irreversible impairment of kidney function. It has been proposed that among the probable causes of CKDnt would be chronic exposure to pesticides, working conditions with exposure to high temperatures and dehydration, chronic use of anti-inflammatory drugs, high consumption of alcohol and tobacco. These risk factors also contribute with the high morbidity and mortality of CKD, since exposure increases with the diagnoses advanced-stage CKD [5,6].

The Pan-American Health Organization defined CKDnt as impaired kidney function with a glomerular filtration rate (GFR) below 60 mL/min/m^2^ [7]. in absence of predisposing factors for traditional CKD (namely, type 2 diabetes mellitus, essential hypertension, heart disease, urinary tract malformations, immunological and congenital diseases. In addition, the diagnosis of CKDnt includes kidney damage defined by structural abnormalities (i.e., renal atrophy without obstructive pattern) or abnormality in the urinary sediment as a marker of kidney damage. There may be exposure to occupational risk factors or living in a risk area [8].

In Panama, since 2014, an increase in patients who meet the criteria for CKDnt have been reported in the provinces of Coclé, Herrera, and Los Santos [9]. In 2017, Panamanian health authorities started the Information System for the Epidemiological Surveillance of CKD to establish mandatory notification of all patients diagnosed with CKD in all its stages and to identify associated risk factors [10]. The system aims to diagnose and establish the real prevalence of CKD in Panama to inform public policies for promotion, prevention, and treatment of CKD. The 2017 Preventive Health Census of the Ministry of Health estimated the national prevalence of CKD at 3.24% [11].

The objective of this study is to characterize the risk factors and clinical phenotype of patients diagnosed with CKDnt from the provinces of Herrera and Los Santos in the central region of Panama and to compare them with patients with CKDt from the same geographic area.

## Methods

We conducted a retrospective, descriptive study with the patients records at the Nephrology Department of the Dr. Gustavo N. Collado Hospital. This hospital is the nephrology reference center for Central Panama (provinces of Herrera and Los Santos, serving a population estimated in 2012 at 214,539 inhabitants) [12].

Records between January 1 and December 31, 2018 and a minimum of two evaluations by Nephrology at least 3 months apart during the study period were included. Records also reported at least two urinalyses and blood samples. The patient record was also the data source for sociodemographic variables, personal history, laboratory results, and renal ultrasound evaluation.

Data was extracted into MS Excel (The Microsoft Corporation; Redmond, WA) and the clean dataset was exported to Stata v. 11.0 (StataCorp, LLC; College Station, TX) for data analyses. Analyses included descriptive statistics. Fisher’s exact test was used to compare proportions and the Mann-Whitney U test to compare medians, setting alpha at 0.05 for statistical significance when comparing the frequencies of the CKDt and CKDnt groups.

## Results

The chart review identified 224 records of patients diagnosed with stage 3 and 4 CKD during 2018. After two reviews of the inclusion criteria, 106 patient files were included (Figure 1).

**Figure 1.**
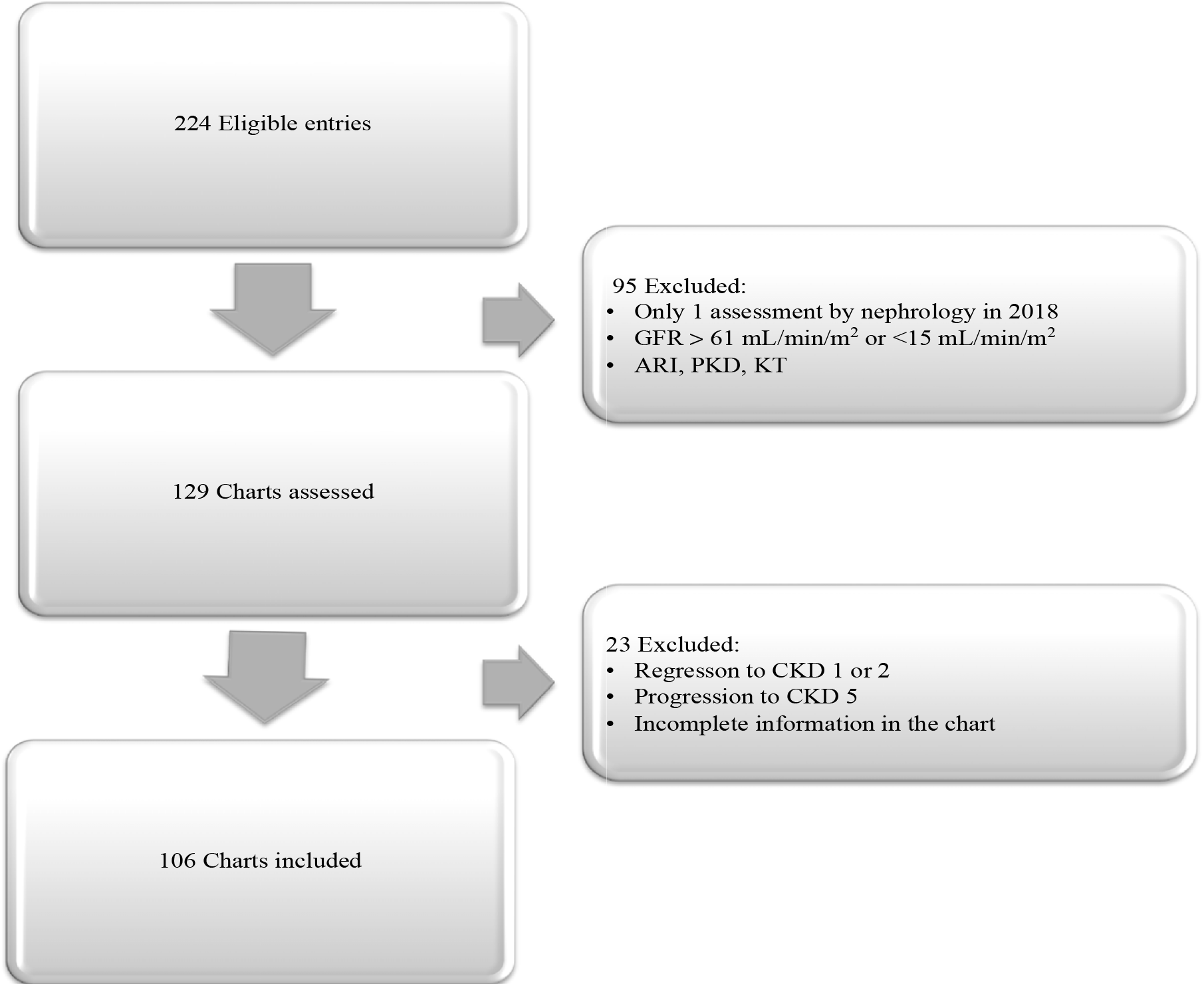
Patient chart selection flow diagram. ARI: acute renal insufficiency; CKD: chronic kidney disease; GFR: glomerular filtration rate; KT: kidney transplant; PKD: polycystic kidney disease.

Of the 106 patient charts included, 66% (70) were male and 34% (36) were female, with a median age of 68.8 years. Forty-five percent of patients were 70 years or older. The median weight was 73 kg, the median height was 1.61 m and the median body mass index (BMI) was 27.7 kg/m^2^.

In the occupation evaluation, 22% of the patients were retired, followed by 20% agricultural workers, 19% household administrators, 14% unemployed, and 13% practiced other professions. Eighty percent of the patients had a history of essential hypertension, 30% type 2 diabetes mellitus, 19% cardiovascular disease, 15% hyperuricemia, and 10% cerebrovascular disease.

Of the total of 106 patients included with a diagnosis of CKD, 14% (*n*=15) met the definition of CKDnt, while 86% (*n*=91) were diagnosed with CKDt. **Table 1** summarizes the relevant clinical, history, ultrasound, and laboratory data for both groups.

**Table 1.** Relevant Clinical and Paraclinical Data of patients with CKDnt and CKDt.

| Characteristics | CKDt (n=91) | CKDnt (n=15) | P value |
| --- | --- | --- | --- |
| Median age, years | 68.8 | 55.4 | <0.001 |
| Median weight, kg | 73.7 | 70.7 | 0.50 |
| Median height, m | 1.6 | 1.64 | 0.14 |
| Median BMI, kg/m<sup>2</sup> | 28.03 | 25.85 | 0.09 |
| Gender |  |  | 0.016 |
| Men | 56 (61) | 14 (93) |  |
| Women | 35 (38) | 1 (7) |  |
| Agricultural workers | 14 (15) | 9 (60) | <0.001 |
| Transportation workers | 0 (0) | 3 (20) | <0.001 |
| Personal history of: |  |  |  |
| Type 2 diabetes mellitus | 31 (34) | 0 (0) | 0.02 |
| Hypertension | 80 (88) | 5 (33) | <0.001 |
| Cardiovascular disease | 20 (22) | 0 (0) | 0.04 |
| Cerebrovascular disease | 11 (12) | 0 (0) | 0.32 |
| Obesity | 1 (1) | 0 (0) | 0.84 |
| CKD | 17 (19) | 2 (13) | 0.80 |
| Obstructive uropathy | 9 (10) | 0 (0) | 0.40 |
| Hyperuricemia | 12 (13) | 4 (27) | 0.37 |
| Renal ultrasound<sup>1</sup> |  |  |  |
| Normal findings | 28 (31) | 2 (13) | 0.16 |
| Renal atrophy | 36 (50) | 13 (87) | <0.001 |
| Hydronephrosis | 7 (10) | 0 (0) | 0.27 |
| Agenesis | 1(1) | 0 (0) | 0.68 |
| Urinalysis |  |  |  |
| Erythrocyturia >3 RBCs/field | 8 (9) | 1 (7) | 0.78 |
| Proteinuria >3+ | 15 (16) | 1 (7) | 0.32 |
| Median blood chemistry values |  |  |  |
| Glucose 1, mg/dL | 105 | 97.5 | 0.07 |
| Glucose 2, mg/dL | 111 | 96.5 | 0.04 |
| Glycated hemoglobin A1c, % | 6.71 | 5.005 | 0.11 |
| Creatinin 1, mg/dL | 1.85 | 2.14 | 0.10 |
| Creatinin 2, mg/dL | 1.89 | 2.17 | 0.13 |
| Blood urea nitrogen, mg/dL | 26.8 | 23.5 | 0.31 |
| Uric acid 1, mg/dL | 6.8 | 7.8 | 0.04 |
| Uric acid 2, mg/dL | 7 | 8.1 | 0.09 |
| Sodium, mEq/L | 139 | 138 | 0.60 |
| Potassium, mEq/L | 4.5 | 4.4 | 0.55 |
| Calcium, mg/dL | 9.75 | 9.7 | 0.72 |
| Triglycerides, mg/dL | 171 | 167 | 0.90 |
| Total cholesterol, mg/dL | 182 | 188 | 0.68 |
| HDL cholesterol, mg/dL | 51.4 | 45.7 | 0.70 |
| LDL cholesterol, md/dL | 108.8 | 101.7 | 0.59 |

<sup>1</sup> Only 72 of the CKDt files had renal ultrasound on file.
BMI: body mass index, CKDt: traditional chronic kidney disease, CKDnt: non-traditional chronic kidney disease, RBC, red blood cells; HDL, high-density lipoprotein; LDL, low-density lipoprotein.

A statistically significant predominance of young men was observed in the group of patients with CKDnt compared to the group with CKDt. Additionally, workers in agriculture or transportation presented with significantly(*P*<0.001) higher frequencies of CKDnt than with CKDt. Although not statistically significant (*P*=0.09), patients with CKDnt tend to exhibit a lower BMI than patients with CKDt. The personal history of type 2 diabetes mellitus, essential hypertension, and cardiovascular disease were significantly less frequent among patients with CKDnt than among those with CKDt. Of note, ultrasound-confirmed that renal atrophy was significantly more frequent (*P*<0.001) among patients with CKDnt than among patients with CKDt. Patients with CKDnt show significantly lower levels of glucose and higher levels of uric acid in the blood.

## Discussion

In Central American countries, the cases of CKDnt have been increasing during the last decades. However, it has not yet been possible to establish a single causal factor, but rather different agents that can produce chronic kidney damage leading to CKDnt. Various causes have been proposed, including chronic occupational exposure to high temperatures and repeated dehydration due to frequent episodes of heat stress, exposure to pesticides with nephrotoxic effect, contamination of water with heavy metals, lifestyle risk factors (e.g., alcoholism, smoking, and diet), all of which could have a significant impact on the pathophysiology of CKD [13–15].

In 2015 in El Salvador, a country where the epidemiology of CKDnt has been extensively studied, the regional prevalence ranged from 7.1% in the Paracentral region; 5.2% in the Eastern region; and 2% in the Central region [16], and that depending on the community and the work activity carried out by the inhabitants, prevalence peaked at 18% in regions near the coast [17]. In Nicaragua, regional prevalence ranged between 8% and 10% [18]. In Panama, a 2014 study estimated the prevalence of CDK requiring dialysis in the province of Coclé at 40 per 100,000 population towards the north of the province, but up to 200 per 100,000 population towards the south of the province. The southern part of Coclé’s economy is characterized by sugar cane and rice agriculture. In this study, the prevalence of CKDnt could not be determined, but it was the first study that reported an increase in CKD prevalence in a specific region of the country [9].

For greater robustness of the data and to confirm chronicity of the renal function, patient charts should have included at least two measurements of serum creatinine instead of only one [19]. Most previously published studies have measured creatinine only once. The diagnosis of CKD required the calculation of the GFR estimate after achieving a stable creatinine value, at least 3 months apart, to establish the degree of chronicity and be able to rule out acute renal failure [20].

In the group of patients with CKDnt, a male predominance was observed compared to the group with CKDt. This has also been observed in other countries, such as El Salvador (78%) [21], Nicaragua (77%) [22] and Costa Rica (70%) [5]. The median age was lower in the CKDnt group (55 years), compared with a median age of 68 years in the CKDt group. The latter is similar to what was observed in the study group of agricultural communities in El Salvador, where the age of the patients was an average of 45 years [21]. The increased frequencies of patients with diagnoses of CKDnt have been observed mainly in some defined areas in Central America. Particularly, areas heavy on agricultural employment are involved, where young labor is hired to perform difficult tasks under the tropical sun. These factors may explain, at least in part, the younger age, lower BMI, and higher prevalence in males presenting with CKDnt. In turn, these patients did not present with chronic diseases typically associated with CKDt, such as type 2 diabetes mellitus and essential hypertension [23].

This study showed that a significantly higher proportion of patients with CKDnt presented with a history of agricultural work, compared to patients in the CKDt group (60% vs. 15%, respectively; *P*<0.001). The central region of Panama has the largest area with crops of corn in the country, with over 26,000 hectares of land per year. The province of Herrera is the fourth in sugarcane production nationwide [24]. The patients in this study belong to the central provinces of Herrera and Los Santos, mostly land at sea level dedicated to agriculture, in a peninsular area that frequently exhibits the highest temperatures in the country of up to 35°C, especially in the dry season between December and April [25]. It has also been described that in the region of Central America and Panama there has been an increase in temperature of up to 1°C between 2010 and 2015 and that this has been important in the central provinces of Herrera and de Los Santos [25]. Predictive models of temperature increase have been proposed, up to 15% in the next decades in the central region of Panama [25]. This could contribute to the increase in cases of CKDnt compared to the CKDt causes in recent years, as a consequence of continuous dehydration due to exposure to high temperatures during strenuous working hours, as has been studied in other groups of sugarcane farmers in Central America [26,27].

Twenty percent of patients with CKDnt worked as drivers of road transport vehicles or heavy equipment. Studies have associated poor diet, continuous hours of work, lack of adequate hydration, and exposure to high temperatures during the sunniest and hottest hours of the day while driving in freight transport drivers [28]. This phenomenon has also been described for workers in a block factory [29]. Thus, this pathology would not be exclusive to agriculture and transportation, but in occupations where there are sustained exposure to high temperatures with risk of dehydration.

Glycosuria without hyperglycemia and HbA1c levels greater than 6.5% can be found in less than 3% of patients with CKDnt [23], probably secondary to the pathophysiological damage caused by CKDnt and not to the presence of type 2 diabetes mellitus [18]. In the present study, no patient in the CKD group presented with type 2 diabetes mellitus, compared to 34% in the CKDt group. A trend towards increased glucose associated with the higher frequency of type 2 diabetes mellitus was observed in the CKDt group, in which poor control could help the progression to worsening of CKDt.

A higher frequency of essential hypertension was evidenced in the group of patients with CKDt compared to the group of patients with CKDnt. Previous studies have found that arterial hypertension is rare in patients with CKDnt and it is highly prevalent in CKDt [30]. Hypertensive nephrosclerosis is a chronic disease that gradually and progressively produces chronic kidney involvement [30,31]. The finding of mild hypertension in some patients with CKDnt (33%) in our study could be due to the fact that it is a consequence of CKD and not the cause, as it has been previously suggested [19]. The presence of mild hypertension of less than 5 years of evolution, associated with advanced chronic kidney disease, in the absence of other risk factors, should lead us to suspect CKDnt.

In this study, uric acid values showed a significant elevation above the normal value in the group of patients with CKDnt compared to CKDt. In dehydration states, fructose is produced endogenously from glucose, leading to uric acid generation, inflammation, and fibrosis in the kidney [32]. Furthermore, asymptomatic hyperuricemia has been associated with the development of mild hypertension, which, when treated and normalized, improves blood pressure control [33 The present study supports the concept that hyperuricemia is a biomarker of CKDnt. More studies are necessary to determine its role in the diagnosis, evolution and prognosis of CKDnt.

In the evaluation of the imaging studies, by renal ultrasound, 87% of the patients diagnosed with CKDnt presented with a decrease in the cortico-medullary relationship with increased echogenicity and renal atrophy. This translates into renal morphological damage observed in early stages of asymptomatic patients. Renal ultrasound studies in El Salvador have found an increase in echogenicity of up to 95% and a decrease in the cortex-medulla ratio in up to 82% of patients with CKDnt [21]. The present study supports the concept that renal atrophy is a clinical marker of CKDnt. More studies are necessary to determine its role in the diagnosis, evolution and prognosis of CKDnt.

Patients with CKD frequently present proteinuria mostly due to glomerular involvement, hypertension, or diabetic nephropathy. In the present study, there were no significant differences proteinuria levels between patients with CKDnt and CKDt. This may be due to the fact that patients included in this study had moderate stages (3 and 4), and not advanced stages of CKD. In patients with CKDnt, low proteinuria (<1 g/dL) was observed. Renal biopsy studies in patients with CKDnt have shown that the histopathological damage is not due to damage to the glomeruli but rather to tubulointerstitial nephropathy [34,35]. This could explain the difference in the presentation of proteinuria that they have these patients in advanced stages of CKD.

There are various studies with patients affected by CKDnt, but there are few prevalence studies with progression to end-stage CKD [23,36]. In the present study, in the group of patients with CKDt, five patients progressed to stage 5 CKD, requiring initiation renal replacement therapy. At follow-up, no patient in the CKDnt group progressed to terminal illness. In early stages of kidney involvement by an external agent, recovery depends on exposure time, as has been observed in drug-caused interstitial nephropathies. When the causative agent is withdrawn early, the patient can recover due to the fact that the damage does not involve the glomeruli. When evaluating histopathological studies, it is important to point out that damage in CKDnt mainly affects the tubules and interstices and not the glomeruli [37,38], so that early identification could delay the progression to terminal disease. Due to the lack of knowledge on prevalence, the information on the frequency of patients diagnosed with CKDnt who progress to renal replacement therapy is still uncertain in Central America [39]. This entity has different names depending on the region, either as CKDnt, CKD of unknown cause, or Mesoamerican nephropathy, which further complicates its epidemiological traceability [15]. Thus, it is essential not only to diagnose it, but also consistently code it as the same diagnosis.

There are environmental exposures that interact with gene function. Several occupational-environmental risk factors predisposing to CKDnt were already discussed, with a possible summative effect and sustained for a certain time that would lead to kidney involvement. However, not all people who live in the same geographic region with the same gender and occupational risk factors will develop kidney involvement leading to CKDnt. Thus, so it is also believed that genetic and epigenetic factors could play an important role in the etiology of CKDnt [40]. It is possible that individual genetic susceptibilities in a triggering environment could induce development and/or worsening of renal involvement [41]. Longitudinal follow-up studies of patients in early stages that could include the genetic and epigenetic component that can also contribute to the identification of biomarkers in patients at risk and help prevent kidney involvement [42].

A limitation of this study is the small sample size in the case of patients with CKDnt. Thus, the interpretation of its implications, including considerations on risk factors, should be prudent. Another limitation is that not all patients had renal ultrasound available to compare the results. Despite its limitations, this study makes important contributions to the understanding of CKDnt since to our knowledge it is the first published study in Panama and one of the few in Central America to address the clinical presentation of patients with early stage CKDnt compared to patients with CKDt.

## Conclusions

CKDnt is more common in younger-aged male patients than in patients with CKDt. CKDnt is diagnosed in areas where agricultural activity predominates; however, it is not the only work activity to which this condition has been associated, but more generally it is related to long-term strenuous work and high temperatures that could lead to dehydration.

Because CKDnt remains asymptomatic for a long time, early detection is important in patients with elevated creatinine values, without comorbidities such as diabetes and hypertension, in which it is important to identify whether there are other risk factors for occupational and environmental exposure and efforts should be directed at halting the progression of the disease at an early stage. The early detection of patients with CKDnt could have an important effect in preventing progression to terminal CKD.

It is important to provide information and education to the population that works in chronic exposure to high temperatures. Likewise, work should be done on the generation of public policies so that employers are aware of the risk factors and allow the necessary rest and hydration times during the working hours of their employees at risk.

New prospective follow-up studies are important to allow the study of genetics, epigenetics, the identification of biomarkers and a better understanding of the pathophysiology, as well as the better therapeutic management of patients at risk or diagnosed with CKDnt.

## Data Availability

All data produced in the present work are contained in the manuscript

## Abbreviations

CKD: chronic kidney disease
CKDnt: nontraditional chronic kidney disease
CKDt: traditional chronic kidney disease
DALYs: Disability-adjusted life years
GFR: glomerular filtration rate
BMI: body mass index

## Acknowledgments

The authors would like to acknowledge Humberto López Castillo, MD, PhD for his review of the manuscript. Iván Landires is a member of the Sistema Nacional de Investigación (SNI), which is supported by Panama’s Secretaría Nacional de Ciencia, Tecnología e Innovación (SENACYT).

## Authors’ contributions

Conceptualization, K.C., N.B., V. N-S., and I.L.; methodology, K.C., N.B., B.H., M.P., C. R., V. N-S., and I.L.; software, K.C., and I.L.; validation, K.C., N.B., V. N-S., and I.L.; formal analysis, K.C., N.B., V. N-S., and I.L.; investigation, K.C., N.B., B.H., M.P., C. R., V. N-S., and I.L.; resources, K.C., V. N-S., and I.L.; data curation, K.C., N.B., B.H., M.P., C. R., V. N-S., and I.L.; writing—original draft preparation, K.C., N.B., V. N-S., and I.L.; writing—review and editing, I.L.; visualization, K.C., N.B., V. N-S., and I.L.; supervision, K.C., and I.L.; project administration, K.C., and I.L.; funding acquisition, K.C., V. N-S., and I.L.; All authors have read and agreed to the published version of the manuscript.

## Funding

No funding was received for this study.

## Availability of data and materials

All data generated or analysed during this study are included in this published article.

## Ethics approval and consent to participate

The study was conducted according to the guidelines of the Declaration of Helsinki, and approved by the Interinstitutional Ethics Committee of the Social Security Fund and the National Directorate for Teaching and Research (DENADOI-SIBI-008-2020). The need for informed consent was waived by the Interinstitutional Ethics Committee of the Social Security Fund and the National Directorate for Teaching and Research (DENADOI-SIBI-008-2020). No further administrative permissions were needed to access the raw data used in this study. The data used in this study were anonymized before use.

## Consent for publication

Not applicable.

## Competing interests

The authors declare that they have no competing interests.

## Notes

### Competing Interest Statement

The authors have declared no competing interest.

### Funding Statement

This study did not receive any funding

### Author Declarations

Interinstitutional Ethics Committee of the Social Security Fund and the National Directorate for Teaching and Research (DENADOI-SIBI-008-2020)

